# Patterns and outcomes of cardiovascular complications among adult patients with chronic kidney diseases attending Benjamin Mkapa Hospital in Dodoma, Tanzania A protocol of a prospective longitudinal study

**DOI:** 10.1101/2023.06.15.23291478

**Authors:** Mohamed Mbalazi, Alphonce Baraka, Alfred Meremo, John Meda

## Abstract

**Introduction:** Chronic Kidney Disease (CKD) affects more than 800 million people globally, the high cardiovascular mortality and morbidity related to CKD result from unique CKD-related risk factors such as anemia and mineral metabolism disturbances and underlying CKD risk factors such as hypertension, diabetes mellitus, and dyslipidemia. The presence of CKD-related structural and functional alterations in the heart and blood vessels increases the risk of hypertension, left ventricular hypertrophy, and heart failure. In advanced CKD stages, the increased risk of hospitalization, heart failure worsening, stroke and death are commonly seen due to increased risk of ischemic heart disease, arrhythmias, and pericarditis.

**Methodology:** The study will follow a prospective longitudinal design recruiting patients 18 and above years with CKD stage 3 confirmed by estimated glomerular filtration rate (eGFR) < 60ml/min/1.73m2 for the past 3 months, or proteinuria of greater than 200mg/g attending Benjamin Mkapa nephrology and cardiology clinics. Cardiovascular complications will be identified following screening .as per operational definition and a comprehensive baseline assessment. A monthly assessment of cardiovascular complications will be done for a period of 6 months through clinical examinations, imaging and laboratory tests. Cornell and Sokolow-Lyon’s ECG criteria will detect LVH, but ECHO will rule it out. The Framingham criteria will be used to diagnose heart failure. A 12-lead ECG will detect arrhythmias, and an ECG plus cardiac markers will be used to diagnose ischemic heart disease. ECG and ECHO findings of pericardial sac thickening or fluid presence will indicate pericarditis. Continuous and discrete variables will be summarized as mean ±standard deviation (SD), medians, and interquartile ranges, while categorical data will be summarized as frequency and proportion. A binomial logistic regression model will be used to assess the association between independent variables and CVD outcomes

## Introduction

Chronic kidney disease is defined as impairment of kidney structure or function for a duration of 3 months that’s defined as a decrease in eGFR of less than 60 mL/min/1.73 m2 or proteinuria of greater than 200 mg/g [1]. Cardiovascular disease is a major public health concern around the world, accounting for 30% of worldwide mortality, equivalent to 17 million fatalities out of a total of 57 million deaths per year, with an alarming 80% of these deaths occurring in (LMICs)[2]. The prevalence of CVD in CKD is 26.8%, 33.4%, 39.1% and 47.2% in Japan, the United States, Spain and the United Kingdom, respectively[3] while in advanced CKD stages, it ranges from 50-80%.[4]. while in a study that was conducted in Tanzania the prevalence of CVD in CKD was 16.2%[5].

The risk of cardiovascular complications among CKD patients is increased due to several factors such as hypertension, diabetes mellitus, smoking, high levels of cholesterol or triglycerides, anaemia, chronic inflammation and endothelial dysfunction[6–10] factors such as hypertension, diabetes mellitus, smoking, high levels of cholesterol or triglycerides, anaemia, chronic inflammation and endothelial dysfunction[6–10] In the early stages of CKD (stages 1-3), the most common forms of CVD are hypertension peripheral artery disease, and left ventricular hypertrophy[12]. As CKD progresses to peripheral artery disease, and left ventricular hypertrophy[12]. As CKD progresses to peripheral artery disease, and left ventricular hypertrophy[12]. As CKD progresses to mortality,[14]. This highlights the importance of close monitoring and effective management mortality,[14]. This highlights the importance of close monitoring and effective management cardiovascular complications among CKD patients include heart attack, stroke, worsening heart failure, and death.[16,17] In addition, patients with CKD and cardiovascular disease may experience a decline in quality of life and increased healthcare costs[18].

Patients with CKD need to be closely monitored for cardiovascular complications and to receive appropriate treatment to reduce their risk of developing cardiovascular complications[19]. This may include lifestyle changes, medication, and close monitoring of cardiovascular risk factors such as blood pressure, lipid levels, and blood sugar levels[20]. Therefore, this study aims to determine the prevalence, pattern and outcomes of cardiovascular complications among adult patients with chronic kidney disease in Dodoma, cardiovascular complications among adult patients with chronic kidney disease in Dodoma, Tanzania

## Materials and methods

### Study aims

1. To determine the prevalence of cardiovascular complications among patients with chronic kidney disease attending Benjamin Mkapa Hospital in Dodoma Tanzania.
2. To determine the pattern of cardiovascular complications among patients with chronic kidney disease attending Benjamin Mkapa Hospital in Dodoma Tanzania.
3. To determine the outcomes of cardiovascular complications among patients with chronic kidney disease attending Benjamin Mkapa Hospital in Dodoma Tanzania.

### Study design

A prospective longitudinal observational study will be conducted among adult patients with CKD attending clinics at medical department BMH from May 2023 to June 2024.

### Study area

Dodoma is Tanzania’s capital city located in the eastern-central region; The BMH is the Zonal top referral hospital, as well as the teaching hospital for the University of Dodoma. It offers a range of services, including dialysis, renal transplant, diagnostic coronary angiography, and percutaneous coronary intervention. The hospital serves around 11.3 million population according to the 2022 Tanzania National Census [21]. The Bed capacity at the Benjamin Mkapa Hospital is around 400. Patients are brought in from lower-level hospitals in the region as well as from neighboring regions such as Singida, Manyara, Tabora, Morogoro, and Iringa. The Benjamin Mkapa hospital has an average attendance of 18213 patients per month; patients with medical conditions ranging from 2800-3110. About 30% of these patients are from cardiology and nephrology clinics.

### Sample size estimation

To estimate the sample size, the proportionate formula used in prospective cohort studies will be used[22]

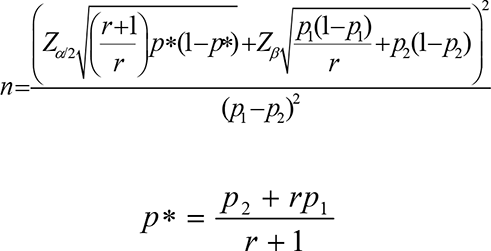

Where by

r = ratio between the two groups

p1 = CV-Complication prevalence (obtained from literature)

p2 = CV-Complications six-moth prevalence observed or expected from the study

p1 – p2 = effect size

*Z*_*β*_ = standard normal variate for statistical power *Z*_∝/2_= standard normal variate for the significance level

The prevalence of cardiovascular complications among CKD patients (based on the study done at Muhimbili -Tanzania is 16.2%[5]

The expected prevalence in this study is expected to be 25%

Therefore;

r = 1

p1 = 16.2%

p2 = 25%

p1 – p2 = -8.8%

*Z*_*β*_ = 0.84 for statistical power of 80%

*Z*_∝/2_ = 2.56 for a significance level of 99%

N= Sample size.

Considering the 20 % attrition rate in our setting[6]

Therefore, the minimum sample size estimated is 234 patients.

### Inclusion criteria

i. Patients more than 18 years of age.
ii. All patients who attend the medical department for at least 6 months with features of chronic kidney disease (stage 3-4) with or without CVD (left ventricular hypertrophy heart failure, pericarditis, ischaemic heart disease and arrhythmia)
iii. Those who consented will be invited to the study.

### Exclusion criteria

i. Patients with a history of or current malignancy, liver disease, or chronic lung disease confirmed from their units because they will exacerbate cardiovascular outcomes and pause a challenge during follow-up.
ii. Female patients who are pregnant or planning to become pregnant during the study period

### Sampling procedure

The sample will be obtained using the consecutive random sampling approach, which involves choosing every patient who will meet the inclusion criteria upon admission or attendance at the nephrology department until the desired sample size is reached. All patients who have visited or been admitted to Benjamin Mkapa Hospital, who has been attending a nephrology clinic for more than 6 months, and who have an eGFR of less than 60 ml/min/1.73 m3 or proteinuria greater than 200 mg/g, will be enrolled after giving their informed consent, or a proxy consent from a close family member.

#### Primary outcome

The outcomes of interest are left ventricular hypertrophy, heart failure, pericarditis, ischaemic heart disease and arrhythmias.

**Left ventricular hypertrophy** will be diagnosed through 12-lead ECG criteria, a Sokolow-Lyon (sum of S wave in V1 and R wave in V5-6 > 35 mm) and Cornell (sum of R wave in aVL and S wave in V3 > 28 mm in men, > 20 mm in women). Because of the low sensitivity of both above-mentioned criteria, an ECHO will be used to assess LVMI (>110 g/m2 in women, >125 g/m2 in men) or LVWT (>11 mm) for LVH diagnosis)[23].

**Heart failure** the diagnosis of heart failure will be established based on Framingham criteria that include clinical characteristics and chest x-ray findings, further characterization of heart failure types either heart failure with reduced, mildly reduced or preserved ejection fraction will be done based on echocardiogram findings[24]

**Arrhythmias** will be diagnosed by a 12-lead electrocardiogram (ECG) machine (model C12G, manufactured by ART Technology Medical Co. Ltd 2019). Supraventricular arrhythmias include sinus tachycardia (HR > 100 bpm, narrow QRS), sinus bradycardia (HR < 60 bpm, wide P-R interval), atrial flutter (saw-toothed wave, 2:1 ratio in lead II, III, aVF), and atrial fibrillation (absence of P-wave, irregular R-R interval, often seen in V1). Individuals who will present with palpitations, shortness of breath, dizziness, anxiety, and chest discomfort despite a normal resting ECG will be subjected to a 24-hour Holter ECG to detect transient or intermittent paroxysmal arrhythmias. Ventricular arrhythmias consist of ventricular tachycardia (absence of P-wave, regular wide QRS) and ventricular fibrillation (no P-wave, irregular and narrow QRS)[25,26]

.**Pericarditis** will be diagnosed using a physical examination, 12 lead ECG and ECHO.

**Ischaemic heart disease** patients with symptoms and traditional risk factors for stable ischemic heart disease will be subjected to stress tests [27]. Patients will be stratified using the Duke Treadmill score. Patients with high risk (less than -10) and intermediate risk (-10 to +4) scoring more than 10 on the Framingham risk score will be candidates for coronary angiography.

#### Secondary outcomes

##### Worsening of heart failure

Will be defined as chronic heart failure symptoms that deteriorate despite previous stable background treatment necessitating an urgent need for an emergency visit, hospitalization, escalation of therapy, outpatient IV diuretic therapy, or outpatient oral therapy[4,28].

##### Stroke

The risk of stroke is higher in people who have cardiovascular diseases such as heart failure, high blood pressure, or atrial fibrillation[23,29]. Stroke as per the World Health Organization is defined as "rapid development of clinical signs of focal or global disturbance of cerebral function lasting more than 24 hours or leading to death, with no apparent cause other than vascular origin" [30,31]. To confirm the stroke diagnosis, these patients will receive a SIEMENS (SOMATOM Definition Flash) non-contrast brain CT scan. Additionally, within the first 14 days following the stroke, patients with stroke-like symptoms but negative CT scans for haemorrhagic stroke and unknown ischemic stroke status will be recruited for a study-specific MRI brain scan[32].

#### Mortality

**All-cause mortality** will refer to the overall mortality rate, including death from any cause. It includes deaths caused by cardiovascular events (such as heart failure, arrhythmia, ischaemic heart disease or strokes), as well as deaths resulting from other causes, such as infections, cancer, accidents, or any other health conditions[16].

**Cardiovascular death** will refer to deaths caused by cardiovascular complications such as failure, arrhythmia, ischaemic heart disease or stroke that necessitate admission rehospitalizations or sudden cardiac arrest[7].

#### Study variables

##### Aim 1 study variables

These variables will address the prevalence of cardiovascular complications among patients with CKD. This will help to know the burden of cardiovascular complications among CKD patients.

##### Aim 2 study variables

These variables will determine the pattern of a specific cardiovascular complication seen more frequently among patients with CKD

##### Aim 3 study variables

The variable will address the outcomes of cardiovascular complications at month 6 assessing the worsening heart failure, stroke, development of arrhythmias and death.

#### Participant characteristics

All adults over the age of 18 who have been receiving care at Benjamin Mkapa Hospital’s nephrology clinic for more than six months and have been diagnosed with CKD stage 3–4 with proteinuria will be included in the study. Both men and women inpatient and outpatient are included, and their cardiovascular complications will be evaluated through a clinical examination, laboratory results, ECHO, and ECG, read by the principal investigator and verified by an experienced cardiologist at the cardiology clinic.

#### Data collection process

##### Evaluation of participants

A standardized questionnaire developed through the available evidence will be tested on a small number of participants. After being tested and improved, participants who consent will be interviewed, and baseline characteristics including age, gender, height, weight, BMI, blood pressure, and pulse will be acquired. Furthermore, an assessment of cardiovascular complications (LVH, Heart failure, pericarditis, IHD, and arrhythmias) will be captured.

#### Clinical variable measurements

##### Weight

The patient will be asked to wear light clothing and no shoes. The weight will be measured using a Seca weighing scale made in German (2012) that is calibrated in kilograms. The patient will stand on the scale with their feet slightly apart and their weight evenly distributed.

##### Height

To measure height accurately, the patient will be asked to stand barefoot with their back straight against a flat surface of a stadiometer. The head, shoulders, buttocks, and heels are kept in contact with the surface while the patient is looking straight ahead. The height is then measured from the top of the head to the feet by using a vertical measuring rod with a sliding horizontal headpiece. The Seca 213stadiometer from German will be used and height will be recorded in meters.

##### Body mass index

The National Health Service (NHS-UK) BMI calculator will be used to compute the body mass index (BMI)[33].

##### Radial pulse assessment

Two operators will assess the radial pulse. "Operator 1" will use a stethoscope to listen to the patient’s heart sounds, and "Operator 2" will palpate the patient’s radial pulse while they are seated. Once everyone operator is ready, a timer will be used to count for one minute, the procedure will be repeated one more time and recorded. The two operators will alternate roles on various patients to remove prejudice. The apex-pulse deficit will be calculated from the difference between the rates counted by the two operators at the end of one minute. A deficit of 10 or more is suggestive of atrial fibrillation with a sensitivity and specificity of 62.8% and 85.7%, respectively [34].

##### Blood pressure (BP)

According to the 2018 AHA/ACC Hypertension guidelines for standard measurement of BP, blood pressure will be recorded by an automated digital machine (AD Medical Inc. brand)[35]. A patient will be considered to have hypertension if the blood pressure is greater than 140/90 millimetres of mercury (mmHg) after three measurements taken at least five minutes apart, or having a history of hypertension or a history of taking antihypertensive medication[36].

##### Left ventricular hypertrophy

Through an ECG Sokolow-Lyon voltage criteria with a sensitivity and specificity of 32% and 100% respectively will be used to define LVH. This criterion is based on the summation of S wave V1 and R wave in V5-6. LVH is considered when the sum exceeds 35 mm. Another method is through Cornell voltage criteria; summation of the amplitude of the R wave in lead aVL and the amplitude of the S wave in lead V3. LVH will be considered when the sum exceeds 28 mm in men and 20 mm in women, the sensitivity and specificity are 41% and 98% respectively[37]. Because of the low sensitivity of both above-mentioned criteria, an ECHO will also be used to pick the ventricular hypertrophy by assessing Left ventricular mass index (LVMI) or left ventricular wall thickness (LVWT). LVH will be defined as LVMI greater than 110 g/m2 in women and 125 g/m2 in men or LVWT greater than 11 mm [38].

##### Heart failure

The clinical Framingham criteria for the diagnosis of HF will be used, a patient must have at least two major criteria or one major criterion and two minor criteria[24].

**Table 1.**
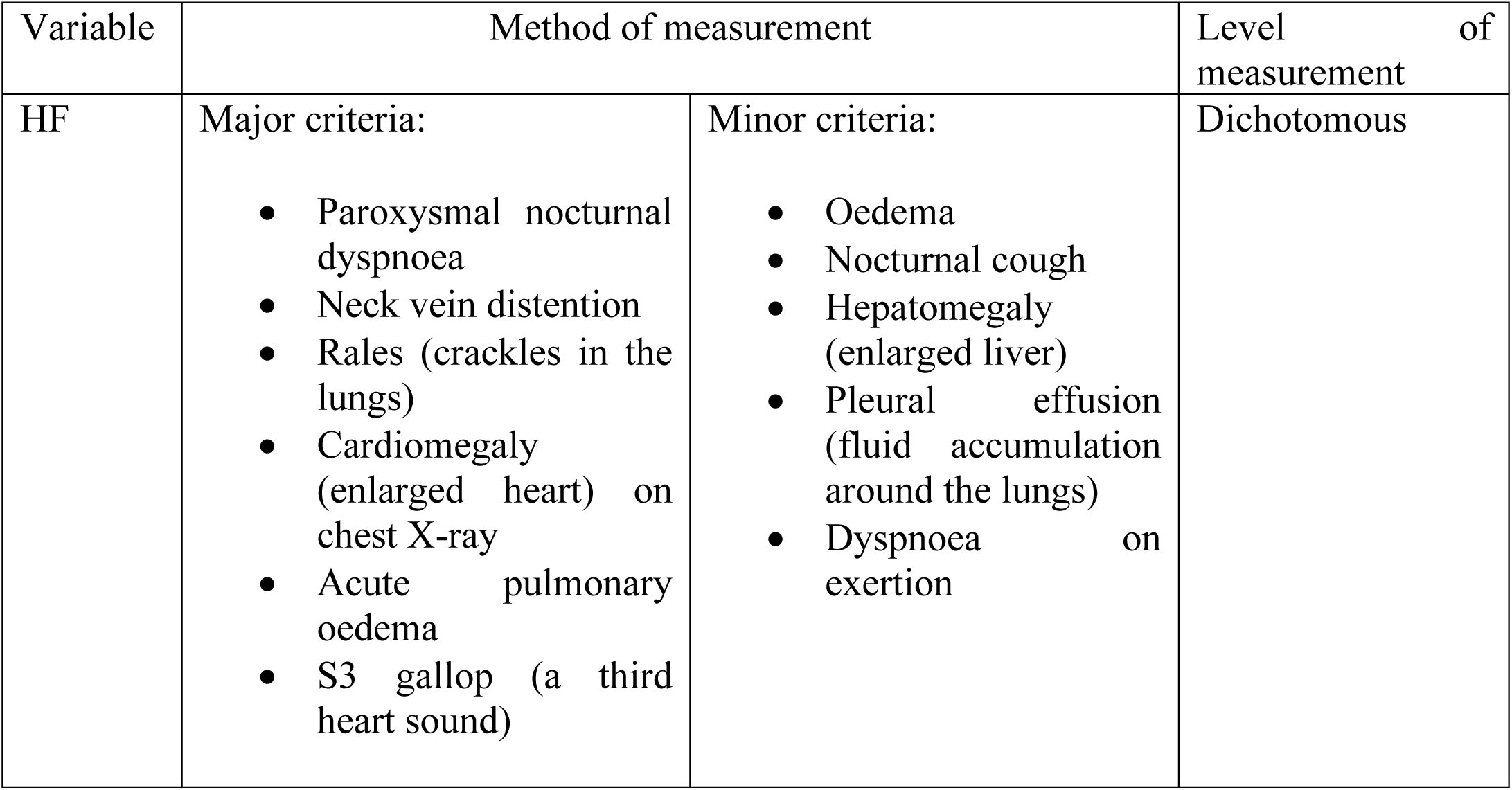
Criteria for clinical diagnosis of heart failure.

Again, an ECHO will assess ejection fraction (EF), heart failure patients will be grouped into three major categories; heart failure with reduced ejection fraction (EF less than 40%), heart failure with mildly reduced ejection fraction (EF between 41 and 49) and heart failure with preserved ejection fraction (EF more than 50%) [39].

##### Arrhythmia

ECG machine (model C12G, manufactured by ART Technology Medical Co. Ltd 2019) will be utilized to assess cardiac arrhythmias, of which supraventricular arrhythmias will refer to abnormal heart rhythms that originate above the ventricles including; sinus tachycardia (HR>100bpm, narrow QRS, narrow P-R Interval, presence of P-wave before every QRS, upright P wave in lead II and downward in aVR), sinus bradycardia (HR<60, P-wave before every QRS, downward in aVR wide P-R interval), atrial flutter (regular saw-toothed wave in lead II, III, aVF with a ratio of 2:1), atrial fibrillation (absence of a P-wave before every QRS, irregular R-R wave interval, mostly seen in V_1_). Ventricular arrhythmias, on the other hand, will include ventricular tachycardia (absence of P-wave, regular wide QRS) and ventricular fibrillation. (no P-wave, irregular and narrow QRS) [25,26,40].

##### Ischemic heart disease

Ischemic heart disease encompasses both unstable ischaemic heart disease (USIHD) and stable ischaemic heart disease (SIHD). Unstable ischaemic heart disease will be diagnosed in an individual who presents clinically with chest pain that occurs at rest, increases in frequency or severity, or lasts longer than stable angina pain, often radiating to the left arm, neck, or jaw[41,42]Based on ECG, ST-elevation myocardial infarction (STEMI) will be defined as ST-segment elevation more than 1 millimetre in two or more contiguous leads (STEMI), whereas ST-segment depression, T-wave inversion, or non-specific T wave changes will be considered as (NSTEMI/unstable angina). Cardiac biomarkers, particularly troponin I of more than 0.05ng/ml with a sensitivity of 98% and specificity of 100% for myocardial infarction will confirm acute myocardial infarction and differentiate NTEMI and unstable angina [42,43]

Stable ischemic heart disease (SIHD) is patients with chest pain that are predictable and usually triggered by physical exertion or emotional stress, lasting for a few minutes and relieved by rest or medication[44]. In the background of traditional risk factors for ischemic heart disease such as obesity, hypertension, or diabetes[45]. Framingham risk score will be used to stratify patients[43]. Those who will score less than 10% and their 12-lead ECG will show signs of previous heart damage by displaying (Q-wave, S-T and T-wave changes) will be low-risk patients and will continue with medical management (aspirin, statin and b-blockers) while those who will score more than 10 and 20 will be categorized as intermediate and high-risk patients who will need treadmill stress ECG for further risk stratification[46]. A stress test will measure how the heart responds to physical stress, patients with a Duke Treadmill score of +5 or more are considered low risk and have 1% mortality risk per year and they don’t need further assessment. Candidates with a score of less than 4 will be considered as an intermediate and high-risk group eligible for coronary angiography[47–49]

**Table 2.**
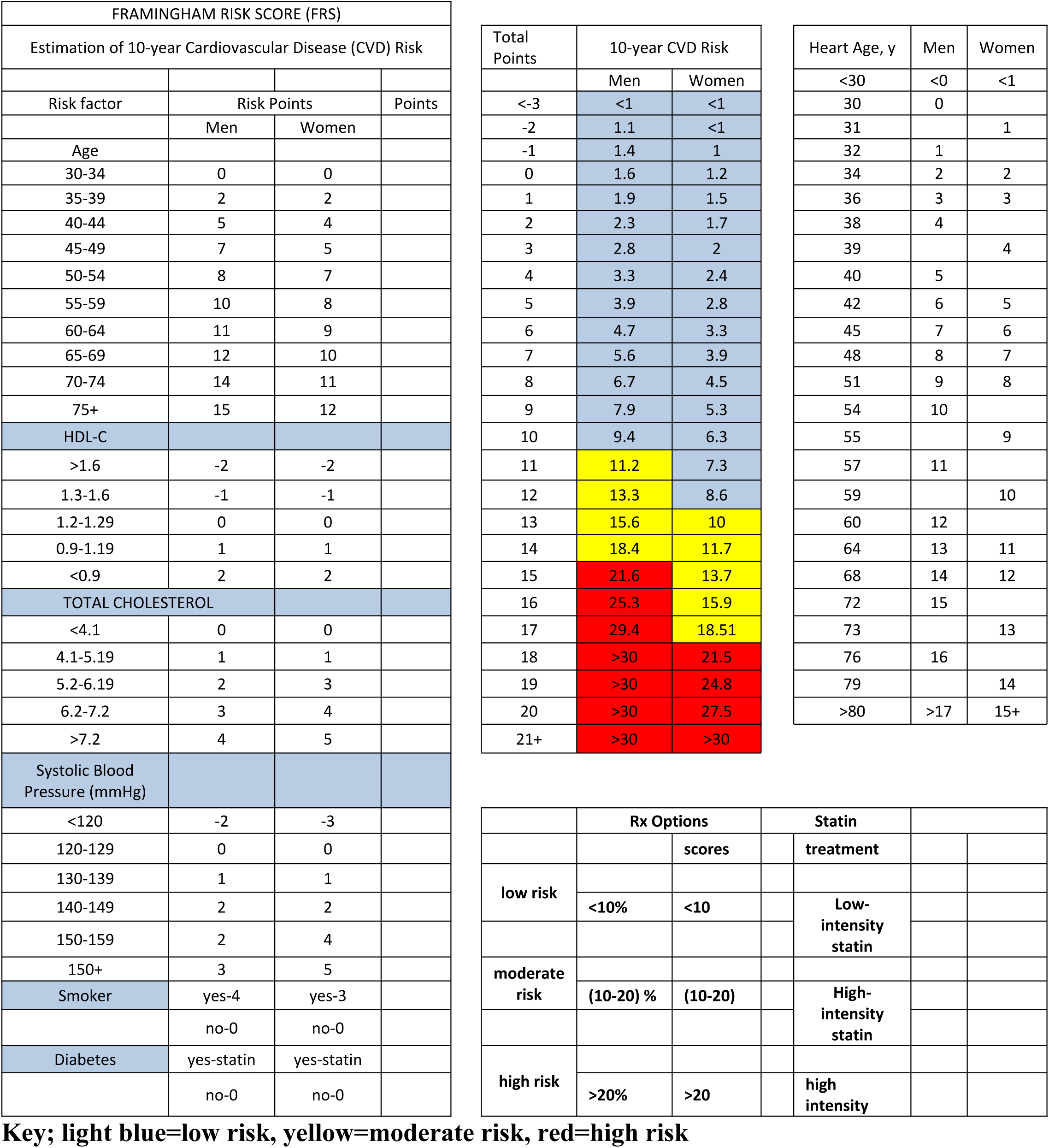
Risk stratification for cardiovascular disease: Framingham Risk Score. .[50]

**Table 3:**
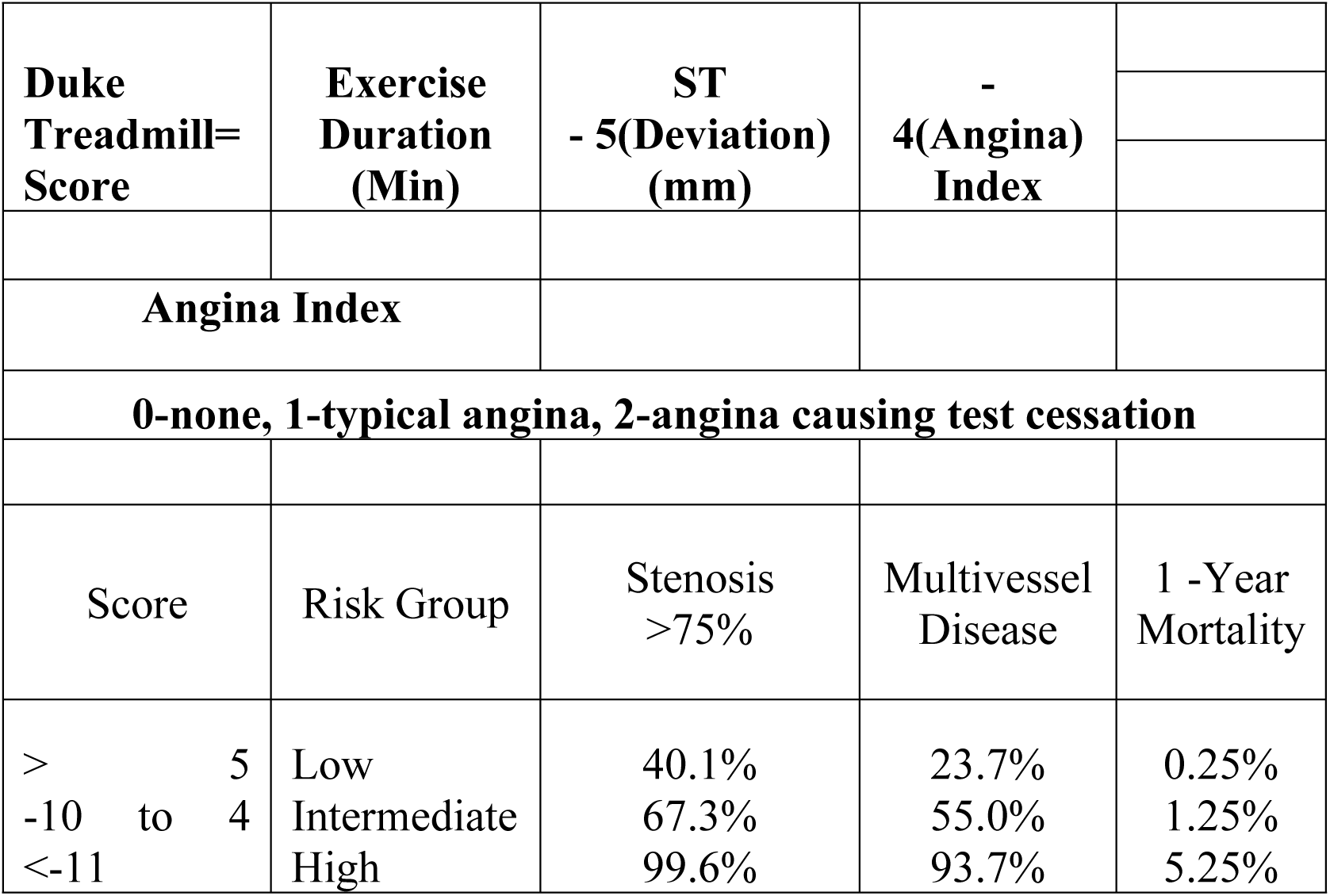
Duke treadmill score-Risk stratification for patients with IHD[51].

| Duke<br>Treadmill=<br>Score | Exercise<br>Duration<br>(Min) | ST<br>- 5(Deviation)<br>(mm) | -<br>4(Angina)<br>Index |  |
| --- | --- | --- | --- | --- |
| **Angina Index** |  |  |  |  |
| **0-none, 1-typical angina, 2-angina causing test cessation** |  |  |  |  |
| Score | Risk Group | Stenosis<br>>75% | Multivessel<br>Disease | 1 -Year<br>Mortality |
| > 5 | Low | 40.1% | 23.7% | 0.25% |
| -10 to 4 | Intermediate | 67.3% | 55.0% | 1.25% |
| <-11 | High | 99.6% | 93.7% | 5.25% |
| Duke Treadmill= Score | Exercise Duration (Min) | ST - 5(Deviation) (mm) | - 4(Angina) Index |  |
| Angina Index |  |  |  |  |
| 0-none, 1-typical angina, 2-angina causing test cessation |  |  |  |  |
| Score | Risk Group | Stenosis >75% | Multivessel Disease | 1 -Year Mortality |
| > 5 | Low | 40.1% | 23.7% | 0.25% |
| -10 to 4 | Intermediate | 67.3% | 55.0% | 1.25% |
| <-11 | High | 99.6% | 93.7% | 5.25% |

##### Pericarditis

The diagnosis of pericarditis will be reached by the presence of diffuse ST segment elevation in all precordial leads on 12 lead ECG or thickening or presence of fluid in the pericardial sac on echocardiogram[52]. However, in a patient with CKD, diagnosis of pericarditis can be challenging, as the symptoms overlap with those of other conditions that are common in CKD patients, such as fluid overload and heart failure[53]. Therefore, a comprehensive evaluation of the patient’s medical history, physical examination and imaging studies is necessary to diagnose pericarditis in a patient with CKD [54].

#### Laboratory and imaging investigations

##### Sample collections and processing

The procedure for sample collection will be explained to the participant, and a lab order form will be reviewed to ensure that all of the needed sample criteria are met. 3 inches from the cubital fossa, a tourniquet will be applied for 60 seconds. A 70% alcohol wipe will be used to clean the venepuncture site, and a 10 cc syringe will be used to draw blood for laboratory tests; blood sample in a yellow caped tube for lipid profile, urine protein creatinine ratio (UPCR), serum creatinine and blood urea nitrogen (BUN), electrolytes (potassium, sodium, calcium and chloride), and troponin I. Centrifugation at 300 rpm for 5 minutes to separate the serum from the whole blood will be done, two aliquots are made: one for the lipid profile and the other for the serum electrolytes, UPCR, serum creatinine/BUN and troponin. However, for samples anticipated to be completed within 2 hours, samples will be stored at 2 to 8 degrees Celsius. Clinical chemistry automatic analyser Erba machine XL-180 with serial number 160239 from Germany will be used to analyse the sample. The following values will be used as normal reference ranges for blood chemistry Calcium (1.12-1.14) mmol/l, potassium (3.5-5.1) mmol/l, sodium (136-145) mmol/l, and chloride (98-107) mmol/l are the electrolytes[55]. FBG (7 mmol/l) and RBG (11.1 mmol/l) are the blood glucose levels[56]. Total cholesterol is 5.17 mmol/l, triglycerides are 2 mmol/l, LDL is 4.13 mmol/l, and HDL is 1.03 mmol/l for men and 1.28 mmol/l for women[57]. Urea 30-40 mg/dl, serum creatinine 0.6-1.5 mg/dl[58], UPCR 200mg/g[59] and troponin 0.04ng/ml[60].

##### Random blood glucose

By the sample collection manual SM-1-03.3 of the BMH laboratory, a blood sample is taken from a finger prick to assay blood sugar. With the candidate’s permission, the palm is turned up and the index, ring, or middle fingertip is selected. Pressure is then administered to the fingertip to enhance blood flow. After cleaning the fingertip with methylated spirit 70% from centre to edge, let it dry for 15 minutes. The finger will be held below the level of the elbow and a fresh, sterile lancet is firmly pressed against the tip to pierce the skin and promote blood flow. The blood sample will be analysed by ACCU-CHECK Active Roche glucometer machine. The participant will be given a ball of cotton wool to hold on the finger for 10 minutes to stop bleeding after the sample has been taken, and the lancet is then disposed of in a Sharp disposal box. According to the American Diabetes Association, hyperglycaemia is defined as random blood sugar levels that are higher than 11.1, or fasting levels that are higher than 7. Diabetes is diagnosed when there is a fasting blood sugar level of 7., random blood glucose levels that are higher than 11.1, and symptoms of hyperglycaemia, or glycated haemoglobin levels that are higher than 6.5%[9].

#### Echocardiography

The patient will be asked to remove their clothing from the waist up and put on a gown and lie on their left side. Electrodes will be placed on their chest to monitor their heart rate and rhythm during the test. A gel will be applied to the patient’s chest and an echocardiogram will be done by a cardiologist. A transducer will be placed on the patient’s chest to obtain the images. Patients may be asked to breathe in and hold their breath at certain times to help get clear images. Image interpretation will be verified by another cardiologist for verification. An ECHO machine (model Vivid ^TM^ T9 manufactured by GE Healthcare, USA 2018) will assess the anatomy and function of the heart by using echocardiography variables; ischemic heart disease (regional wall motion abnormalities-kinesia), pericarditis (pericardial thickening and Pericardial effusion) LVH (Intra-ventricular septum, posterior wall diameter of >11mm). Systolic dysfunction of the left ventricle (EF less than 50%), diastolic dysfunction (mitral inflow velocity more than4cm, mitral valve annular velocity lateral fewer than10cm/s, or septal less than 7cm/s, TR Velocity more than 2.8m/s, LA volume index more than 34ml/m^2^) congestive heart failure (ventricular wall hyperkinesia)[61].

#### Electrocardiography

I will make sure the patient is comfortable and relaxed and explain the procedure to the patient then the patient will be undressed from the waist up, and ensure their chest is clean and dry. Patients will be in a supine position with their arms at their sides and ensure no movement or talk during the test. A 12 lead ECG (model C12G, manufactured by ART Technology Medical Co. Ltd 2019) will be used. Electrodes will be placed as follows: V1: 4th intercostal space to the right of the sternum, V2: 4th intercostal space to the left of the sternum, V3: Midway between V2 and V4, V4: 5th intercostal space in the midclavicular line, V5: Anterior axillary line at the same level as V4 and V6: Midaxillary line at the same level as V4 and V5. Red colours limb lead RA: Right arm, yellow coloured limb lead LA: Left arm Black coloured limb lead RL: Right leg and Green coloured limb lead LL: Left leg. After recording, the ECG will be analysed and interpreted with the help of two experienced cardiologists.

### Data management and analysis

Data will be collected and stored in an Excel spreadsheet on an encrypted computer. The data will be converted to IBM SPSS PC version 25 and then evaluated and cleaned. Continuous variables will be presented as mean ±standard deviation (SD), medians, and interquartile ranges while categorical data will be presented as frequency or proportional. The chi-square test will be utilized to establish the association between two categorical variables, such as sex, gender, BMI, BP, UPCR, Cholesterol level, Hemoglobin level, Diabetes mellitus, CKD stages, and the presence of LVH, HF, pericarditis, IHD, and arrhythmia.

The univalent logistic regression model will be used to analyze the association between independent variables and cardiovascular complications. The selection of variables for multivariate analysis will include variables with a p-value less than 0.2 in univariate analysis. Adjusted odds ratios (a OR) will be reported, along with their 99% confidence intervals (CI). Variables with p value less than 0.01 will be considered statistically significant.

### Ethical consideration

Ethical approval was sought from the University of Dodoma’s Director of Research and Publications, with reference number MA.84/261/02/’A’/59/17. Following that, with reference number AB.150/293/01/296, ethical approval for data collection at the Benjamin Mkapa Hospital was acquired. Before enrolling, participants will be briefed about the study and ensure that they understand what the research is all about. They will then choose whether or not to participate in the study, and those who accept will sign a written informed consent. The confidentiality of participants will be protected by assigning them numbers.

### Study Timeline

To meet the estimated sample size, the study is anticipated to take 12 months the first participants will be recruited on May 2023, and each patient will require a 6-month follow-up.

## Discussion

CKD patients have a higher prevalence of cardiovascular complications around 50% compared to the general population 26%[4]. The most common forms of CVD are hypertension, peripheral artery disease, and left ventricular hypertrophy[13]. As CKD progresses to advanced stages (stages 4-5), the incidence of ischemic heart disease and heart failure increases[13] In advanced stages of CKD, CVD is a major contributor to morbidity and mortality[14]. This highlights the importance of close monitoring and effective management of CVD risk factors in CKD patients, especially in advanced stages.[15]. The outcomes of cardiovascular complications among CKD patients include heart attack, stroke, worsening heart failure, and death. Cardiovascular problems in CKD patients can result in a heart attack, stroke, heart failure, or death. Given the scarcity of data in Sub-Saharan Africa, this study has the potential to give valuable information on the burden, trends, and outcomes of cardiovascular complications among CKD patients in our environment. However, as a prospective cohort research, maintaining touch with all cohort participants can be difficult, especially if the cohort is big and the follow-up time is protracted. Before publication, the final results will be submitted to the University of Dodoma library and the study site, Benjamin Mkapa Hospital, and a manuscript will be created for submission to peer-reviewed publications.

## Data Availability

No datasets were generated or analysed during the current study. All relevant data from this study will be made available upon study completion.

## Acknowledgement

I would like to express my gratitude to my supervisors, Dr Alfred Meremo, Dr John Meda and Dr Alphonce Baraka for their invaluable guidance and support throughout the development of this study protocol. Their expertise and insight have been instrumental in shaping the research questions and methodology. I am grateful for their unwavering commitment to ensuring the rigour and validity of this study and their willingness to provide constructive feedback and encouragement at every step of the way. Without their guidance, this protocol would not have been possible.

## Author Contributions

**Conceptualization**: Mohamed Mbalazi, Alfred Meremo, John Meda

**Data curation**: Mohamed Mbalazi

**Formal analysis**: Mohamed Mbalazi

**Investigation**: John Meda

**Methodology**: Mohamed Mbalazi, Alphonce Baraka, Alfred Meremo, John Meda.

**Supervision**: Alphonce Baraka, Alfred Meremo, John Meda

**Writing - original draft**: Mohamed Mbalazi

**Writing- review & editing**: Alphonce Baraka, Alfred Meremo, John Meda

